# Identifying core MRI sequences for reliable automatic brain metastasis segmentation

**DOI:** 10.1101/2023.05.02.23289342

**Authors:** Josef A Buchner, Jan C Peeken, Lucas Etzel, Ivan Ezhov, Michael Mayinger, Sebastian M Christ, Thomas B Brunner, Andrea Wittig, Björn Menze, Claus Zimmer, Bernhard Meyer, Matthias Guckenberger, Nicolaus Andratschke, Rami A El Shafie, Jürgen Debus, Susanne Rogers, Oliver Riesterer, Katrin Schulze, Horst J Feldmann, Oliver Blanck, Constantinos Zamboglou, Konstantinos Ferentinos, Angelika Bilger, Anca L Grosu, Robert Wolff, Jan S Kirschke, Kerstin A Eitz, Stephanie E Combs, Denise Bernhardt, Daniel Rückert, Marie Piraud, Benedikt Wiestler, Florian Kofler

## Abstract

**Background:** Many automatic approaches to brain tumor segmentation employ multiple magnetic resonance imaging (MRI) sequences. The goal of this project was to compare different combinations of input sequences to determine which MRI sequences are needed for effective automated brain metastasis (BM) segmentation.

**Methods:** We analyzed preoperative imaging (T1-weighted sequence ± contrast-enhancement (T1/T1-CE), T2-weighted sequence (T2), and T2 fluid-attenuated inversion recovery (T2-FLAIR) sequence) from 339 patients with BMs from six centers. A baseline 3D U-Net with all four sequences and six U-Nets with plausible sequence combinations (T1-CE, T1, T2-FLAIR, T1-CE+T2-FLAIR, T1-CE+T1+T2-FLAIR, T1-CE+T1) were trained on 239 patients from two centers and subsequently tested on an external cohort of 100 patients from five centers.

**Results:** The model based on T1-CE alone achieved the best segmentation performance for BM segmentation with a median Dice similarity coefficient (DSC) of 0.96. Models trained without T1-CE performed worse (T1-only: DSC = 0.70 and T2-FLAIR-only: DSC = 0.73). For edema segmentation, models that included both T1-CE and T2-FLAIR performed best (DSC = 0.93), while the remaining four models without simultaneous inclusion of these both sequences reached a median DSC of 0.81-0.89.

**Conclusions:** A T1-CE-only protocol suffices for the segmentation of BMs. The combination of T1-CE and T2-FLAIR is important for edema segmentation. Missing either T1-CE or T2-FLAIR decreases performance. These findings may improve imaging routines by omitting unnecessary sequences, thus allowing for faster procedures in daily clinical practice while enabling optimal neural network-based target definitions.

## Introduction

Brain metastasis (BM) delineation is a time-consuming process in clinical practice and research alike. Automated BM segmentation algorithms can be used to assist in this task. They require only a fraction of the time an experienced clinician needs to perform delineation while achieving an overlap with the reference segmentation within the range of interrater variability [1,2].

We have previously developed a model for the simultaneous segmentation of both contrast-enhancing BMs and surrounding T2-weighted fluid-attenuated inversion recovery (T2-FLAIR) hyperintense edema [1]. Like many other approaches to brain tumor segmentation, such as the BraTS challenge [3] or FeTS [4], our model uses four magnetic resonance imaging (MRI) sequences as input, namely a T1-weighted sequence (T1), a T1-weighted sequence with contrast enhancement (T1-CE), a T2-weighted sequence (T2) and T2-FLAIR.

Using fewer input sequences is clearly advantageous. In clinical practice, individual sequences may not be of the required quality, e.g., due to motion artifacts [5]. Furthermore, while a complete brain imaging protocol averages a scan time of about 21 minutes [6], an adapted protocol with only two sequences can decrease duration by about ten minutes. Also, shorter scan times are, in turn, known to reduce patient motion [5]. In addition, using fewer sequences reduces the amount of data that needs to be processed. This results in faster pre-processing times and leaner neural networks.

Although the administration of MRI contrast agents generally results in fewer and less severe adverse effects than the use of iodine-based computed tomography contrast agents, there are still some adverse reactions including rare, life-threatening anaphylactoid reactions [7]. Their use should therefore be carefully considered. Nevertheless, contrast-enhanced sequences are part of many imaging routines, such as in the radiation therapy planning of brain tumors [8]. Thus, BM segmentation algorithms that work without contrast would be of great use.

While some authors have built neural networks for BM segmentation using only T1-CE, they focused only on the BM itself without the surrounding T2-FLAIR hyperintense edema [9,10]. While edema segmentation currently has no relevance for the radiotherapy (RT) planning of BMs, it can be relevant for glioma [11]. Moreover, the delineation of edema may provide valuable information for downstream analysis with techniques such as radiomics [12] or neural network-based feature extraction.

This project aimed to compare neural networks with different combinations of input sequences for the segmentation of the contrast-enhancing *metastasis* and the surrounding T2-FLAIR hyperintense *edema*. All neural networks were tested in a multicenter international external test cohort composed of 100 patients from five different centers to investigate the contribution of different MRI sequences to the segmentation of contrast-enhancing BMs and their surrounding edema.

## Methods

### Automatic segmentation of brain metastases

In our previous work, we focused on how to improve the detection and segmentation of BMs [1]. This project aimed to quantify the contribution of individual MRI sequences to the quality of segmentation. In the following, we will refer to our previous publication and highlight the changes in our workflow.

### AURORA study

Data were collected as part of the *A Multicenter Analysis of Stereotactic Radiotherapy to the Resection Cavity of Brain Metastases* (AURORA) retrospective study conducted by the *Radiosurgery and Stereotactic Radiotherapy Working Group* of the *German Society for Radiation Oncology* (DEGRO) [13]. Inclusion criteria were a resected BM with a known primary tumor and stereotactic RT with radiation dose > 5 Gy per fraction. Exclusion criteria were an interval between surgery and RT > 100 days, premature discontinuation of the RT, and any previous cranial RT. Synchronous unresected BMs were allowed but had to be treated concurrently with stereotactic RT [1]. Institutional ethical approval was obtained (main approval at the Technical University of Munich: 119/19 S-SR; 466/16 S). While the study focuses clinically on the postoperative situation, we analyzed only the preoperative imaging.

### Dataset

We received data from 481 patients from seven centers in total (TUM: Klinikum rechts der Isar of the Technical University of Munich, USZ: University Hospital Zurich, FD: General Hospital Fulda, FFM: Saphir Radiochirurgie/University Hospital Frankfurt, FR: University Hospital Freiburg, HD: Heidelberg University Hospital, KSA: Kantonsspital Aarau). As an extension of the previous study, an additional center was included in the test group (FR).

We analyzed preoperative MRI scans only. For our established preprocessing workflow, we needed four MRI sequences from each patient: T1, T1-CE, T2, and T2-FLAIR.

Unlike in our last workflow [1], only the T2 sequence was allowed to be missing because it was not available for a large fraction of the cohort. If other sequences besides the T2 sequence or multiple sequences were missing, the patient was excluded.

The required sequences were available in sufficient quality for a total of 339 patients (70% of total). We divided the patients into a training cohort of 239 patients from two centers (TUM and USZ) and a test cohort of 100 patients from five centers (FD, FFM, FR, HD, and KSA).

### Data preprocessing

We used the same established preprocessing workflow as previously [1]. In short, we used BraTS-Toolkit [14] to generate co-registered, skull-stripped sequences with an isotropic resolution of 1 millimeter in BraTS space.

A total of 123 T2 sequences were missing (106 (44%) in the training cohort and 17 (17%) in the test cohort). These were synthesized with a generative adversarial network (GAN) [15] by feeding the remaining three sequences into the GAN. The synthesized sequences passed visual inspection.

### Annotation

All images were segmented by a doctoral student (JAB) using the open-source software 3D Slicer (version 4.13.0, stable release, https://www.slicer.org/) [16]. Two separate, non-overlapping labels were segmented: The *metastasis* label, consisting of the contrast-enhancing metastasis and necrosis, and the T2-FLAIR hyperintense *edema* label. The segmentations of the test set patients were reviewed by a senior radiation oncologist (JCP).

### Sequence combinations

To reduce the number of models to be trained, we did not train with every possible combination of input sequences, but instead only analyzed clinically plausible combinations by following these considerations: To identify the exact outline of the BM, T1-CE is required [17]. To quantify the added benefit of administering contrast agents, a comparison between T1 and T1-CE may provide further insight. If the main interest is edema, T2-FLAIR may be sufficient. Additional sequences may further improve the quality of segmentations. We did not train a T2-only model to prevent neural networks from receiving only synthetic data from some patients without original data as input. The model trained with all four sequences is referred to as *baseline* for the remainder of this manuscript. Overall, we trained models with the following sequence combinations:

- T1-CE + T1 + T2 + T2-FLAIR (*baseline*)
- T1-CE only
- T1 only
- T2-FLAIR only
- T1-CE + T2-FLAIR
- T1-CE + T1
- T1-CE + T1 + T2-FLAIR

### Neural Network

We kept all training parameters the same as in our previous study [1]. We implemented spatial flips, Gaussian noise, and random affine transformations to augment our training data. As loss function, we chose an equally weighted Dice + Binary Cross Entropy (BCE) loss, as used by Isensee *et al.* [18]. We trained all networks for a total of 500 epochs. The best model was chosen based on the lowest overall loss in the training set.

All models were trained on a workstation equipped with an Intel 9940X CPU combined with two NVIDIA RTX 8000 GPUs using CUDA version 11.4 in conjunction with Pytorch version 1.13.1 [19] and MONAI version 1.1.0 [20].

### Metrics

We calculated the Dice similarity coefficient (DSC) with the Python package pymia [21]. Unless otherwise noted, segmentation metrics were derived from all segmented BMs. We also calculated the metastasis-wise DSC for each BM. To quantify the correlation between BM volume and metastasis-wise DSC, the Spearman correlation coefficient was used. To assess the BM detection performance, we used a pipeline created by Pan *et al.* [22] to determine the F1-score (F1), sensitivity, and precision. The performance of multiple models was compared with the Kruskal-Wallis rank sum test. Numeric and categorial data in the patient cohorts was compared with the Kruskal-Wallis rank sum test and Pearson’s Chi-squared test, respectively.

## Results

The mean number of BMs, patient demographics, and the number of patients with synthesized T2 sequences in each center are shown in Table 1.

**Table 1:** Mean number of brain metastases, patient demographics, and number of synthesized T2 sequences.

| Characteristic | Training-Cohort |  |  | Test-Cohort |  |  |  |  |  |
| --- | --- | --- | --- | --- | --- | --- | --- | --- | --- |
|  | Overall, N = 239<sup>1</sup> | TUM, N = 162<sup>1</sup> | USZ, N = 77<sup>1</sup> | Overall, N = 100<sup>1</sup> | FD, N = 6<sup>1</sup> | FFM, N = 12<sup>1</sup> | FR, N = 16<sup>1</sup> | HD, N = 42<sup>1</sup> | KSA, N = 24<sup>1</sup> |
| **BMs** | **1.4 ± 0.7** | 1.3 ± 0.7 | 1.6 ± 0.8 | **1.4 ± 0.8** | 1.0 ± 0.0 | 2.1 ± 1.7 | 1.1 ± 0.3 | 1.3 ± 0.6 | 1.4 ± 0.7 |
| **Sex** |  |  |  |  |  |  |  |  |  |
| **f** | **117 (49%)** | 79 (49%) | 38 (49%) | **56 (56%)** | 2 (33%) | 8 (67%) | 9 (56%) | 24 (57%) | 13 (54%) |
| **m** | **122 (51%)** | 83 (51%) | 39 (51%) | **44 (44%)** | 4 (67%) | 4 (33%) | 7 (44%) | 18 (43%) | 11 (46%) |
| **Age at RT start** | **62 (53, 71)** | 62 (53, 71) | 62 (54, 69) | **61 (54, 67)** | 62 (57, 64) | 58 (52, 66) | 58 (49, 66) | 61 (57, 65) | 62 (54, 70) |
| **Primary Diagnosis** |  |  |  |  |  |  |  |  |  |
| **NSCLC** | **84 (35%)** | 35 (22%) | 49 (64%) | **40 (40%)** | 4 (67%) | 6 (50%) | 1 (6.2%) | 18 (43%) | 11 (46%) |
| **SCLC** | **1 (0.4%)** | 0 (0%) | 1 (1.3%) | **1 (1.0%)** | 0 (0%) | 0 (0%) | 0 (0%) | 0 (0%) | 1 (4.2%) |
| **Melanoma** | **41 (17%)** | 23 (14%) | 18 (23%) | **10 (10%)** | 1 (17%) | 1 (8.3%) | 1 (6.2%) | 2 (4.8%) | 5 (21%) |
| **RCC** | **11 (4.6%)** | 9 (5.6%) | 2 (2.6%) | **8 (8.0%)** | 0 (0%) | 1 (8.3%) | 2 (12%) | 3 (7.1%) | 2 (8.3%) |
| **Breast** | **34 (14%)** | 33 (20%) | 1 (1.3%) | **19 (19%)** | 0 (0%) | 4 (33%) | 5 (31%) | 8 (19%) | 2 (8.3%) |
| **GI** | **24 (10%)** | 24 (15%) | 0 (0%) | **11 (11%)** | 0 (0%) | 0 (0%) | 4 (25%) | 5 (12%) | 2 (8.3%) |
| **other** | **44 (18%)** | 38 (23%) | 6 (7.8%) | **11 (11%)** | 1 (17%) | 0 (0%) | 3 (19%) | 6 (14%) | 1 (4.2%) |
| **Synthesized T2** | **106 (44%)** | 103 (64%) | 3 (3.9%) | **17 (17%)** | 6 (100%) | 3 (25%) | 8 (50%) | 0 (0%) | 0 (0%) |

<sup>1</sup>Mean ± SD; n (%); Median (IQR)
We split our dataset in a training cohort (TUM: Klinikum rechts der Isar of the Technical University of Munich, USZ: University Hospital Zurich) and a multicenter external test cohort (FD = General Hospital Fulda, FFM: Saphir Radiochirurgie/University Hospital Frankfurt, FR: University Hospital Freiburg, HD: Heidelberg University Hospital, KSA: Kantonsspital Aarau). There were no significant differences in sex and age at the start of the radiotherapy ( $p = 0.8$ and $0.8$ , Pearson's Chi-squared test and Kruskal-Wallis rank sum test) between the seven centers of our dataset. All seven histologies (non-small cell lung carcinoma (NSCLC), small-cell lung carcinoma (SCLC), melanoma, renal cell carcinoma (RCC), breast cancer, gastrointestinal cancer (GI), and others) were present in both the training and the test cohort.

Table 2 summarizes our model evaluation results. Regarding *metastasis* segmentation, all models that included T1-CE in their selected sequences showed similar performance, with a small but significant difference (median DSC = 0.93-0.96, p < 0.001). In contrast, the models trained only on T2-FLAIR and only on T1 reached a significantly lower median DSC for the *metastasis* of 0.73 (IQR = 0.54-0.84) and 0.70 (IQR = 0.46-0.81). The models trained only on T1-CE or T1-CE and T1 performed even better than *baseline* with a median DSC of 0.96 and 0.95, respectively.

**Table 2:** Volumetric segmentation performance of our selected models.

| Model | Metastasis<sup>1</sup> | Edema<sup>1</sup> |
| --- | --- | --- |
| Baseline | 0.90 \| 0.94 (0.89, 0.95) | 0.88 \| 0.93 (0.86, 0.96) |
| T2-FLAIR | 0.66 \| 0.73 (0.54, 0.84) | 0.78 \| 0.87 (0.71, 0.94) |
| T1-CE + T2-FLAIR | 0.88 \| 0.94 (0.91, 0.96) | 0.85 \| 0.93 (0.84, 0.96) |
| T1-CE | 0.91 \| 0.96 (0.93, 0.97) | 0.82 \| 0.87 (0.79, 0.91) |
| T1-CE + T1 | 0.92 \| 0.95 (0.93, 0.96) | 0.83 \| 0.89 (0.79, 0.92) |
| T1-CE + T1 + T2-FLAIR | 0.89 \| 0.93 (0.88, 0.95) | 0.88 \| 0.93 (0.87, 0.96) |
| T1 | 0.61 \| 0.70 (0.46, 0.81) | 0.73 \| 0.81 (0.66, 0.87) |

<sup>1</sup>Mean | Median (IQR)
We report the mean and median Dice similarity coefficient (DSC) of each model for the metastasis and edema labels. All models including T1-CE perform similarly for metastasis segmentation with a median DSC of 0.93-0.96 with the T1-CE-only model performing best. Adding more sequences does not improve the segmentation quality. The simultaneous presence of T1-CE and T2-FLAIR is important for edema segmentation.

To determine the relationship between BM size and segmentation performance, we divided BMs into three equal groups according to their size: small, medium, and large BMs with a median volume of 1.16, 7.15, and 25.68 milliliters, respectively. The median metastasis-wise DSC in each group is shown in Supplementary Table 1. Our T1-CE-only model achieved a median metastasis-wise DSC of 0.92, 0.96 and 0.95 in the three groups. In total, five BMs of 135 total BMs (3.7%) were missed by the T1-CE-only model. Two examples are shown in Supplementary Figure 1. There was a weak to moderate correlation between metastasis-wise DSC and volume for this model with a Spearman correlation coefficient of 0.32. The *baseline* model again performed worse than the T1-CE model, with the largest difference seen in the small BMs. The T1-only and T2-FLAIR-only models both struggled to segment small BMs, achieving a median metastasis-wise DSC of 0.00 and 0.15, respectively, in this group.

For *edema* segmentation, all models which included both T1-CE and T2-FLAIR (*baseline*, T1-CE + T2-FLAIR, T1-CE + T1 + T2-FLAIR) performed best with a median DSC of 0.93. The remaining three models with only one of these two sequences (T1-CE-only, T2-FLAIR-only, T1-CE + T1) reached a median DSC of 0.87-0.89. Again, the T1-only model performed worst with a median DSC of 0.81 (IQR = 0.66-0.87). A segmentation of the *metastasis* and *edema* generated by our T1-CE-only model is shown in Figure 1.

**Figure 1:**
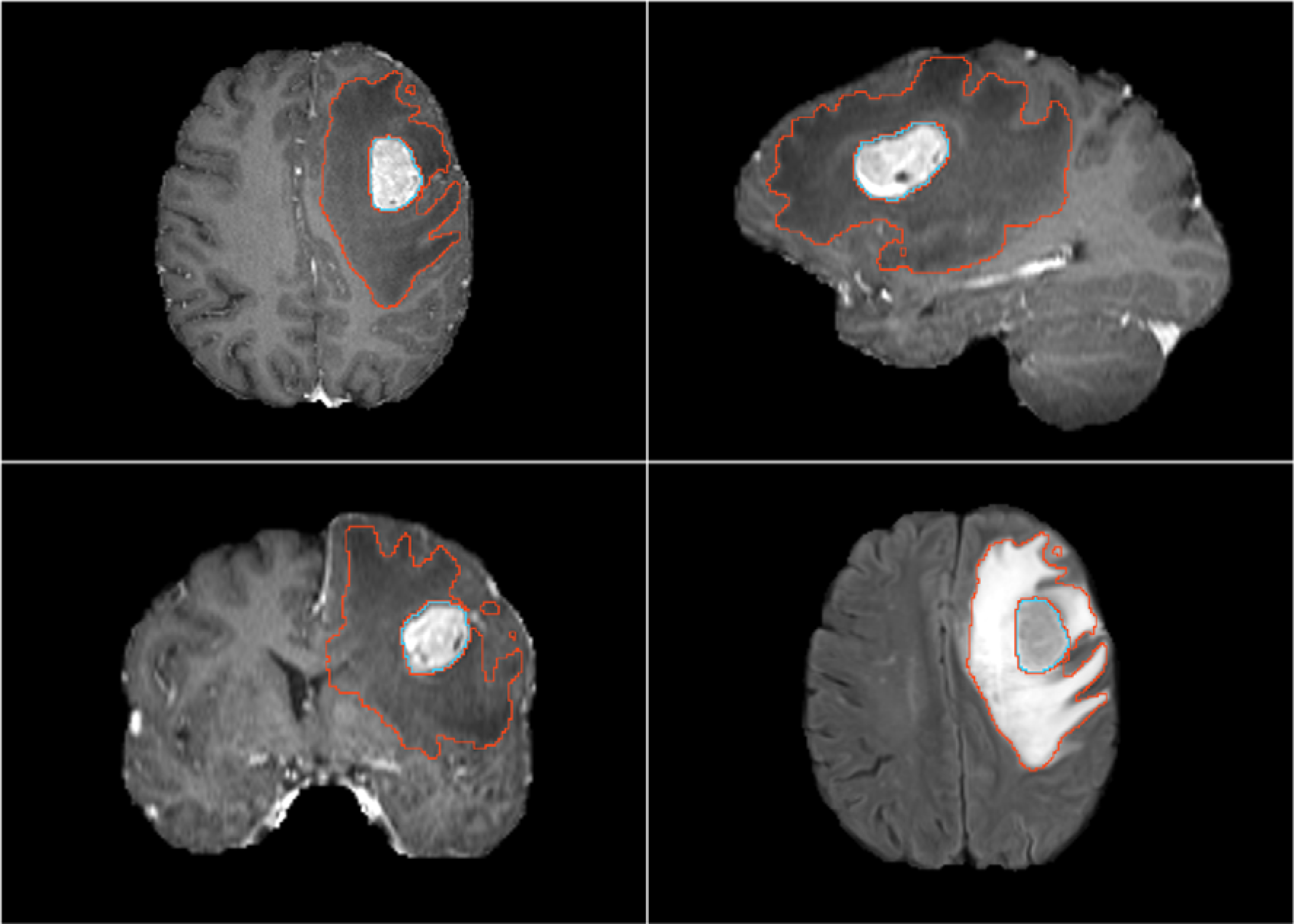
Example of automatic segmentation by our T1-CE-only model. A segmentation of the *metastasis* (in blue) and the *edema* (in red) is shown. Even though the network only received the T1-CE sequence as input (shown in axial, sagittal, and coronal orientation in the top left, top right, and bottom left, respectively), the edema has also been correctly segmented as illustrated by the axial view of the T2-FLAIR (bottom right).

When evaluating the *metastasis* and *edema* labels as a combined *whole lesion* label, the T2-FLAIR-only model exhibited only minimally worse performance than the T1-CE + T2-FLAIR model with median DSCs of 0.94 and 0.95, respectively. This demonstrates that the boundary between *metastasis* and *edema* rather than the outline of the *whole lesion* poses a challenge to the T2-FLAIR-only model. The segmentation metrics for the *whole lesion* label for all models are summarized in Supplementary Table 2. Qualitative inspection of the segmentations supports this thesis (see Supplementary Figure 2).

To check the generalizability of the models, the performance in the individual centers of the test set was compared. As an example, the performance of our T1-CE + T2-FLAIR model for the *metastasis* and *edema* labels is shown in Figure 2 for each center separately. No significant differences were found between the centers. The median DSC for the T1-CE-only model for the metastasis ranged from 0.94 (FFM and FR) to 0.96 (FD, HD and KSA). Excluding the 17 patients with synthetic T2 showed largely similar results: If there was any change at all, it was a slight change in DSC of 0.01-0.02. In the baseline model, the only model that included the T2 sequences among the selected sequences, there was no change in median DSC for *metastasis* and *whole lesion* and an increase from 0.93 to 0.94 for *edema*. The segmentation performance of all 83 patients with four available sequences is shown in Supplementary Table 3.

**Figure 2:**
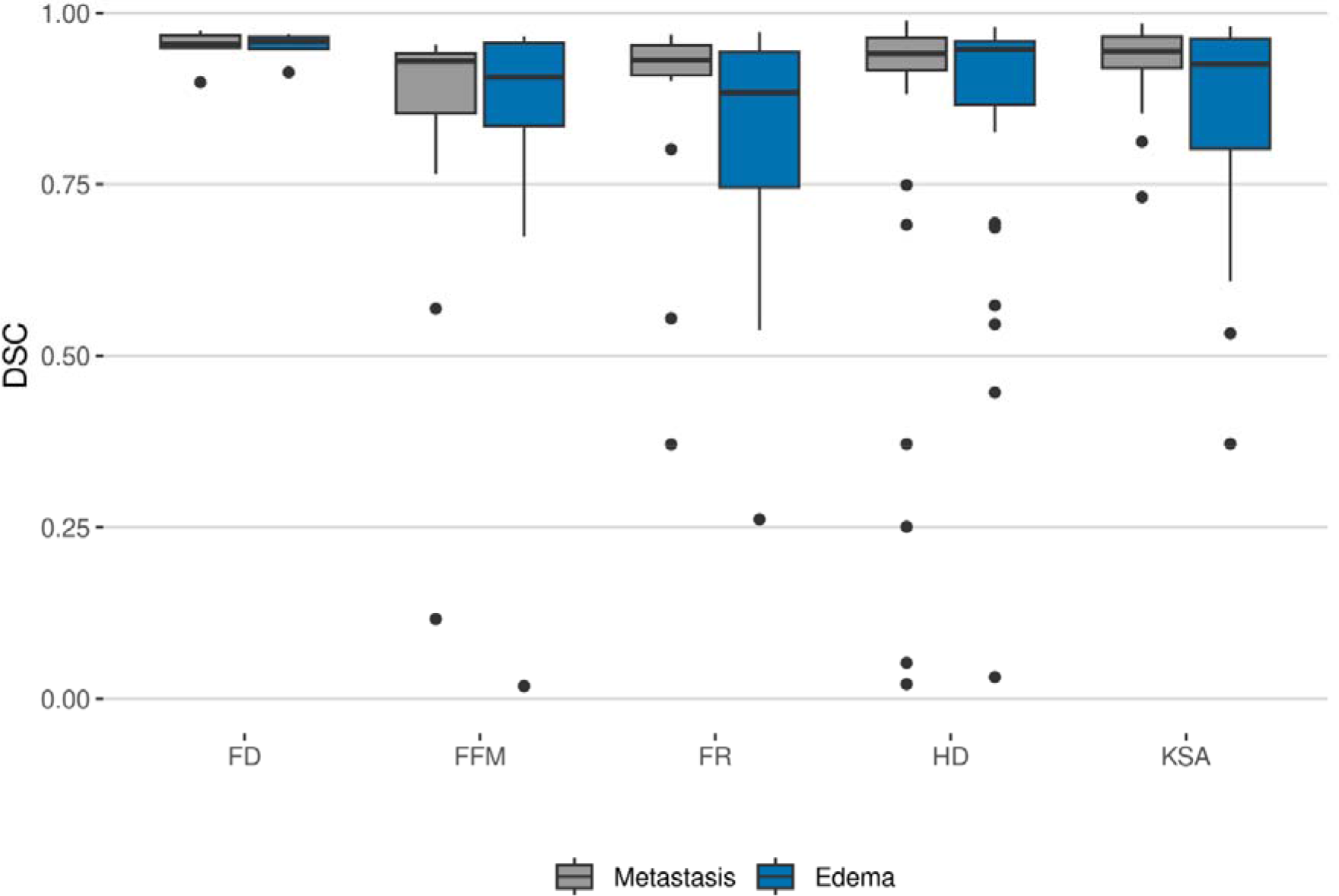
Performance across the five centers of our test set. The segmentation performance of our T1-CE + T2-FLAIR model was stable across all centers as shown by the consistently high Dice similarity coefficient (DSC). There were no significant differences in metastasis and edema segmentation performance (p = 0.094 and 0.6, Kruskal-Wallis rank sum test). While most segmentations were of good quality with a DSC of around 0.9, there were some outliers. This explains the difference between mean and median DSC.

In total, we differentiated between seven different histology groups of primary tumors: non-small cell lung carcinoma (NSCLC), small-cell lung carcinoma (SCLC), melanoma, renal cell carcinoma (RCC), breast cancer, gastrointestinal cancer (GI), and others. Four models (T1-CE, T1-CE + T1, and T1-CE + T1 + T2-FLAIR, T1) showed a significant difference between groups for the segmentation of the BM (p = 0.033, 0.033, 0.013, and 0.047 respectively). The T1-CE-only model reached a DSC between 0.92 (Melanoma) and 0.98 (SCLC). The remaining models performed stable independent of the histology of the primary tumor (Supplementary Table 5).

Table 3 summarizes the BM detection performance. Only mean values are given since the performance was calculated on a per-patient basis and the median performance across all patients was often 1. Patients in our test cohort had 1.4 BMs on average. While all models including T1-CE among their selected sequences showed a high sensitivity of at least 0.96, the T2-FLAIR-only and T1-only models reached only 0.91 and 0.84, respectively. T1-CE-only and T1-CE + T1 detected BMs with a mean precision of 0.97 and 0.92, respectively. In contrast, all models including T2-FLAIR segmented a high number of false positives and therefore achieved a mean precision of only 0.60-0.76. The T1-only model also reached a similar precision of 0.72. As the model with the highest mean number of false positives (1.4), the T2-FLAIR-only model segmented a mean of 2.6 BMs per patient. On the other hand, the T1-CE-only model only labeled 0.08 false positives per patient on average. See Figure 3 for an example patient with five false positives in total segmented by the T2-FLAIR-only model. The F1 showed similar behavior: T1-CE-only and T1-CE + T1 achieved a mean F1 of 0.97 and 0.93 while the remaining models achieved a mean score between 0.66 and 0.81.

**Figure 3:**
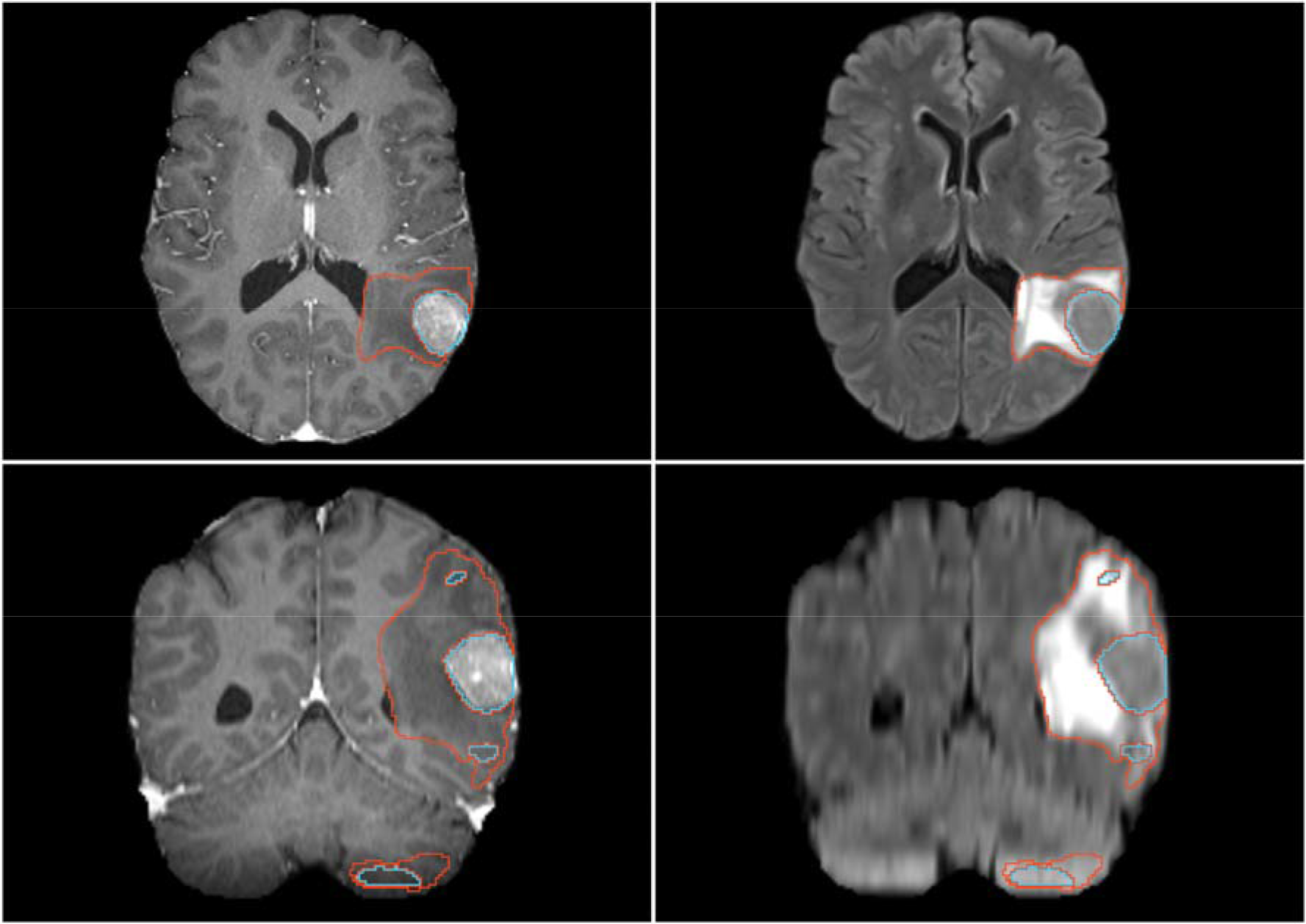
False positives in segmentation of the T2-FLAIR-only model. The segmentation by our T2-FLAIR-only model is shown for a patient with an especially high number of false positives (five in total). On the left, the T1-CE sequence is displayed in the axial and coronal planes. On the right, the same slices of the T2-FLAIR sequence are shown. While the *metastasis* (in blue) and edema (in red) have been correctly identified and labeled, the model also labeled several false positives.

**Table 3:** Metastasis detection performance.

| Model | F1<sup>1</sup> | Sensitivity<sup>1</sup> | Precision<sup>1</sup> |
| --- | --- | --- | --- |
| Baseline | 0.78 ± 0.23 | 0.97 ± 0.10 | 0.72 ± 0.31 |
| T2-FLAIR | 0.66 ± 0.26 | 0.91 ± 0.21 | 0.60 ± 0.32 |
| T1-CE + T2-FLAIR | 0.77 ± 0.24 | 0.97 ± 0.11 | 0.70 ± 0.31 |
| T1-CE | 0.97 ± 0.11 | 0.98 ± 0.08 | 0.97 ± 0.13 |
| T1-CE + T1 | 0.93 ± 0.14 | 0.98 ± 0.09 | 0.92 ± 0.19 |
| T1-CE + T1 + T2-FLAIR | 0.81 ± 0.22 | 0.96 ± 0.12 | 0.76 ± 0.29 |
| T1 | 0.72 ± 0.30 | 0.84 ± 0.32 | 0.72 ± 0.32 |

<sup>1</sup>Mean ± SD

Automatic segmentation from preprocessed files took less than 20 seconds on consumer-grade hardware (NVIDIA RTX 3090), regardless of the model selected.

## Discussion

We compared neural networks with different clinically plausible combinations of input sequences to determine the influence of individual sequences on *metastasis* and *edema* segmentation performance and BM detection performance. For segmenting the *metastasis*, the sole presence of T1-CE is important, and a T1-CE-only protocol appears to be sufficient with a median DSC of 0.96. In contrast, for the segmentation of the *edema*, the combination of T1-CE and T2-FLAIR is crucial, while employing only T2-FLAIR as input leads to worse results. T1-only performed worst in all segmentation tasks. Thus, we consider the administration of contrast agents necessary for BM segmentation.

As with *metastasis* segmentation, the presence of T1-CE was most important for the sensitivity in detecting BMs, and additional sequences did not improve sensitivity. T1-CE-only and T1-CE + T1 detected BMs with the best precision (0.97 and 0.92, respectively). Contrary to expectation, the addition of T2-FLAIR did not improve BM detection performance but instead resulted in more false positives. Together with this article, we are publishing a flexible segmentation tool that chooses the appropriate neural network depending on the availability of sequences.

The weak to moderate correlation between metastasis-wise DSC and BM volume in our T1-CE-only model is due to a slightly worse performance in small BMs. This may be due to the higher proportion of edge voxels in small BMs, which increases the difficulty of segmentation. Nevertheless, the T1-CE-only model achieved a median metastasis-wise DSC of 0.92 in the small BM group (median volume of 1.16 milliliters).

The mean DSC for both the *metastasis* and the *edema* label is on average 0.06 points lower than the median DSC. This shows that while most labels are of very good quality, some outliers reduce the mean. The consistently high performance across the five centers of our test set shows that our models generalize well.

The poor *edema* segmentation performance of our T2-FLAIR-only model might be explained by the way the model generates labels: As in our previous study [1], the model generates an output for the *metastasis* and the *whole lesion*. The *edema* label is then calculated by subtracting the *metastasis* label from the *whole lesion* label to ensure gapless segmentation. Therefore, poor *metastasis* segmentation will also result in a low DSC in the *edema* segmentation.

This new segmentation method has some advantages over our previous workflow: Previously, only one sequence was allowed to be missing or corrupted, which was then synthesized using a generative adversarial network (GAN) [15]. While this allows our previous network to be used on patients with only three available MRI sequences, it adds complexity to the preprocessing workflow. In addition, examinations with multiple missing sequences cannot be segmented with the previous workflow. Furthermore, having to acquire fewer sequences for objective metastasis and edema segmentation benefits both patients and physicians.

To our knowledge, no other publication has performed a comparable in-depth analysis of the contribution of individual MRI sequences to the segmentation performance of metastasis and edema labels. However, for example, Pflüger *et al.* also created a “slim” version of their neural network using only the T1-CE and T2-FLAIR sequences as input in addition to their standard model [23]. They observed a slight but significant decrease in performance for the contrast-enhancing metastasis when using fewer input sequences (median DSC: 0.90 down to 0.89).

Charron *et al.* compared several databases of single MRI sequences (T1-CE, T1, T2-FLAIR) and combinations of them for the detection and segmentation of BMs [24]. They found that when using a single sequence, T1-CE performed best. When two sequences were used, T1-CE + T2-FLAIR resulted in better sensitivity and fewer false positives. The simultaneous use of all three sequences resulted in the best DSC (0.79) and the lowest number of false positives per patient (7.8). These results are only partially comparable to our results because they focused only on metastasis detection and segmentation. In addition, all data were collected from the same center and there was no external or multicentric test cohort. The difference in mean DSC of their best model (T1-CE + T1+ T2-FLAIR) compared to our similar model (0.79 vs. our 0.90) can be partially explained by the higher number and smaller size of BMs in their dataset. The high proportion of edge voxels in small metastases may make segmentation more difficult.

This work has several limitations: The preprocessing workflow we have been using [14] is designed to work with the established four sequences. For our new models to be viable for use in research, preprocessing pipelines must be created that can work with a reduced or variable number of input sequences. The reference annotations were all created by the same person. Thus, the trained neural network adapted the personal segmentation style of our original rater. Even though the segmentations of the test set were checked by an additional rater, a dataset created by multiple raters may hold even greater validity. Because we focused on the imaging of patients who later underwent surgery, many BMs were often larger than metastases that are primarily treated with RT. Especially when trying to detect smaller BMs, T2-FLAIR may be more important than our experiments suggest.

Despite these limitations, we were able to show that neural networks can segment contrast-enhancing BMs as well as their surrounding edemas with a reduced number of input sequences. For the segmentation of BMs, T1-CE-only appears to provide sufficient segmentation quality. For situations, where the edema segmentation is of relevance, such as glioma RT planning, the combination of T1-CE and T2-FLAIR seems to be particularly suitable, as it offers high segmentation performance for both tumor and edema combined with reduced image acquisition time. These findings can help to adapt RT planning MRI protocols and shorten them, thus speeding up procedures in daily clinical practice. Our tool has been uploaded to GitHub and can be accessed via the following link: https://github.com/HelmholtzAI-Consultants-Munich/AURORA.

## Data Availability

The datasets generated and analyzed during the current study are not available.

## Notes

### Competing Interest Statement

The authors have declared no competing interest.

### Funding Statement

This work was funded by the Deutsche Forschungsgemeinschaft (DFG, German
Research Foundation - PE 3303/1-1 (JCP), WI 4936/4-1 (BW)).

### Author Declarations

The ethics committee of Technical University of Munich gave ethical approval for this work (119/19 S-SR; 466/16 S)

### Summary of Updates

An additional center with six patients was added; the results were supplemented with additional evaluations regarding the influence of metastasis size on the quality of segmentation; the influence of the histology of the primary tumor was reviewed

## References

[1] Buchner JA, Kofler F, Etzel L, Mayinger M, Christ SM, Brunner TB, et al. Development and external validation of an MRI-based neural network for brain metastasis segmentation in the AURORA multicenter study. Radiotherapy and Oncology 2023;178:109425. https://doi.org/10.1016/j.radonc.2022.11.014.

[2] Kofler F, Wahle J, Ezhov I, Wagner S, Al-Maskari R, Gryska E, et al. Approaching Peak Ground Truth 2022. https://doi.org/10.48550/arXiv.2301.00243.

[3] Bakas S, Reyes M, Jakab A, Bauer S, Rempfler M, Crimi A, et al. Identifying the Best Machine Learning Algorithms for Brain Tumor Segmentation, Progression Assessment, and Overall Survival Prediction in the BRATS Challenge. ArXiv 2018;10:arXiv:1811.02629. https://doi.org/10.48550/ARXIV.1811.02629.

[4] Pati S, Baid U, Edwards B, Sheller M, Wang SH, Reina GA, et al. Federated learning enables big data for rare cancer boundary detection. Nat Commun 2022;13. https://doi.org/10.1038/S41467-022-33407-5.

[5] Zaitsev M, Maclaren J, Herbst M. Motion artifacts in MRI: A complex problem with many partial solutions. J Magn Reson Imaging 2015;42:887–901. https://doi.org/10.1002/JMRI.24850.

[6] Ellingson BM, Bendszus M, Boxerman J, Barboriak D, Erickson BJ, Smits M, et al. Consensus recommendations for a standardized Brain Tumor Imaging Protocol in clinical trials. Neuro Oncol 2015;17:1188–98. https://doi.org/10.1093/neuonc/nov095.

[7] Hao D, Ai T, Goerner F, Hu X, Runge VM, Tweedle M. MRI contrast agents: Basic chemistry and safety. Journal of Magnetic Resonance Imaging 2012;36:1060–71. https://doi.org/10.1002/jmri.23725.

[8] Popp I, Weber WA, Combs SE, Yuh WTC, Grosu AL. Neuroimaging for Radiation Therapy of Brain Tumors. Topics in Magnetic Resonance Imaging 2019;28:63–71. https://doi.org/10.1097/RMR.0000000000000198.

[9] Zhou Z, Sanders JW, Johnson JM, Gule-Monroe M, Chen M, Briere TM, et al. MetNet: Computer-aided segmentation of brain metastases in post-contrast T1-weighted magnetic resonance imaging. Radiotherapy and Oncology 2020;153:189–96. https://doi.org/10.1016/j.radonc.2020.09.016.

[10] Xue J, Wang B, Ming Y, Liu X, Jiang Z, Wang C, et al. Deep learning-based detection and segmentation-assisted management of brain metastases. Neuro Oncol 2020;22:505–14. https://doi.org/10.1093/neuonc/noz234.

[11] Niyazi M, Andratschke N, Bendszus M, Chalmers AJ, Erridge SC, Galldiks N, et al. ESTRO-EANO guideline on target delineation and radiotherapy details for glioblastoma. Radiotherapy and Oncology 2023;184. https://doi.org/10.1016/j.radonc.2023.109663.

[12] Peeken JC, Wiestler B, Combs SE. Image-Guided Radiooncology: The Potential of Radiomics in Clinical Application. Recent Results in Cancer Research 2020;216:773–94. https://doi.org/10.1007/978-3-030-42618-7_24.

13. AURORA trial – AG Stereotaxie n.d. https://www.degro.org/ag-stereotaxie/projekte/aktuelle-projekte/aurora-trial/ (accessed March 13, 2023).

[14] Kofler F, Berger C, Waldmannstetter D, Lipkova J, Ezhov I, Tetteh G, et al. BraTS Toolkit: Translating BraTS Brain Tumor Segmentation Algorithms Into Clinical and Scientific Practice. Front Neurosci 2020;14. https://doi.org/10.3389/fnins.2020.00125.

[15] Thomas MF, Kofler F, Grundl L, Finck T, Li H, Zimmer C, et al. Improving Automated Glioma Segmentation in Routine Clinical Use Through Artificial Intelligence-Based Replacement of Missing Sequences With Synthetic Magnetic Resonance Imaging Scans. Invest Radiol 2022;57:187–93. https://doi.org/10.1097/RLI.0000000000000828.

[16] Kikinis R, Pieper SD, Vosburgh KG. 3D Slicer: A Platform for Subject-Specific Image Analysis, Visualization, and Clinical Support. Intraoperative Imaging and Image-Guided Therapy 2014:277–89. https://doi.org/10.1007/978-1-4614-7657-3_19.

[17] Sze G, Milano E, Johnson C, Heier L. Detection of brain metastases: comparison of contrast-enhanced MR with unenhanced MR and enhanced CT. AJNR Am J Neuroradiol 1990;11:785.

[18] Isensee F, Jaeger PF, Kohl SAA, Petersen J, Maier-Hein KH. nnU-Net: a self-configuring method for deep learning-based biomedical image segmentation. Nat Methods 2021;18:203–11. https://doi.org/10.1038/s41592-020-01008-z.

[19] Paszke A, Gross S, Massa F, Lerer A, Bradbury J, Chanan G, et al. PyTorch: An Imperative Style, High-Performance Deep Learning Library. Adv Neural Inf Process Syst 2019;32. https://doi.org/10.48550/arXiv.1912.01703.

[20] MONAI Consortium: MONAI: Medical open network for AI (3 2020). https://doi.org/10.5281/zenodo.4323058, https://github.com/Project-MONAI/MONAI n.d.

[21] Jungo A, Scheidegger O, Reyes M, Balsiger F. pymia: A Python package for data handling and evaluation in deep learning-based medical image analysis. Comput Methods Programs Biomed 2021;198. https://doi.org/10.1016/j.cmpb.2020.105796.

[22] Pan C, Schoppe O, Parra-Damas A, Cai R, Todorov MI, Gondi G, et al. Deep Learning Reveals Cancer Metastasis and Therapeutic Antibody Targeting in the Entire Body. Cell 2019;179:1661–1676.e19. https://doi.org/10.1016/J.CELL.2019.11.013.

[23] Pflüger I, Wald T, Isensee F, Schell M, Meredig H, Schlamp K, et al. Automated detection and quantification of brain metastases on clinical MRI data using artificial neural networks. Neurooncol Adv 2022;4. https://doi.org/10.1093/NOAJNL/VDAC138.

[24] Charron O, Lallement A, Jarnet D, Noblet V, Clavier JB, Meyer P. Automatic detection and segmentation of brain metastases on multimodal MR images with a deep convolutional neural network. Comput Biol Med 2018;95:43–54. https://doi.org/10.1016/J.COMPBIOMED.2018.02.004.

