## Supplemental Material for "Identifying core MRI sequences for reliable automatic brain metastasis segmentation"

**Contents:**

- Supplementary Table 1: Relationship between metastasis-wise DSC and BM size
- Supplementary Figure 1: Missed BMs
- Supplementary Table 2: Volumetric segmentation performance of all trained models
- Supplementary Figure 2: Example patient with misidentified boundary metastasis/edema boundary
- Supplementary Table 3: Volumetric segmentation performance for patients with all four sequences
- Supplementary Table 4: Volumetric segmentation performance for the 17 patients with synthetic T2 sequences
- Supplementary Table 5: DSC for metastasis segmentation depending on histology of primary tumor

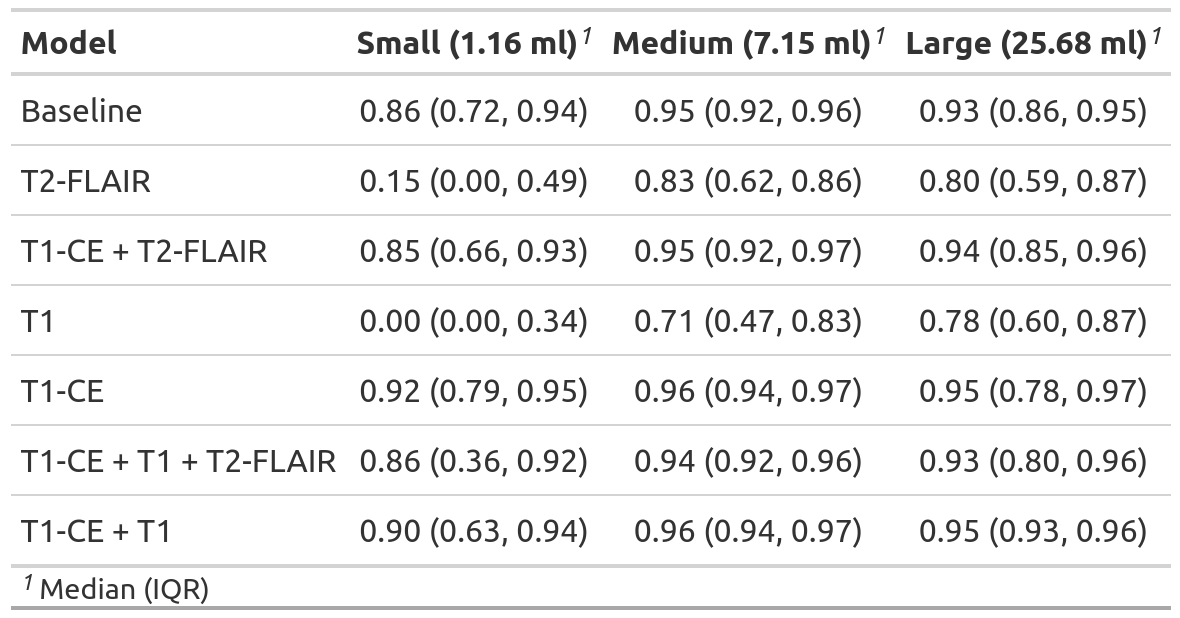

**Supplementary Table 1: Relationship between metastasis-wise DSC and BM size**

The median metastasis-wise DSC for all models is reported for small, medium, and large BMs (median volumes of 1.16, 7.15, and 25.68 milliliters). The T1-CE-only model performed best regardless of the metastasis volume. The lower median metastasis-wise DSC in the small BM group is likely due to a high proportion of edge voxels. The T1-only and T2-FLAIR-only models seem to miss many small BMs and only achieve a median metastasis-wise DSC of 0.00 and 0.15, respectively, in this group.

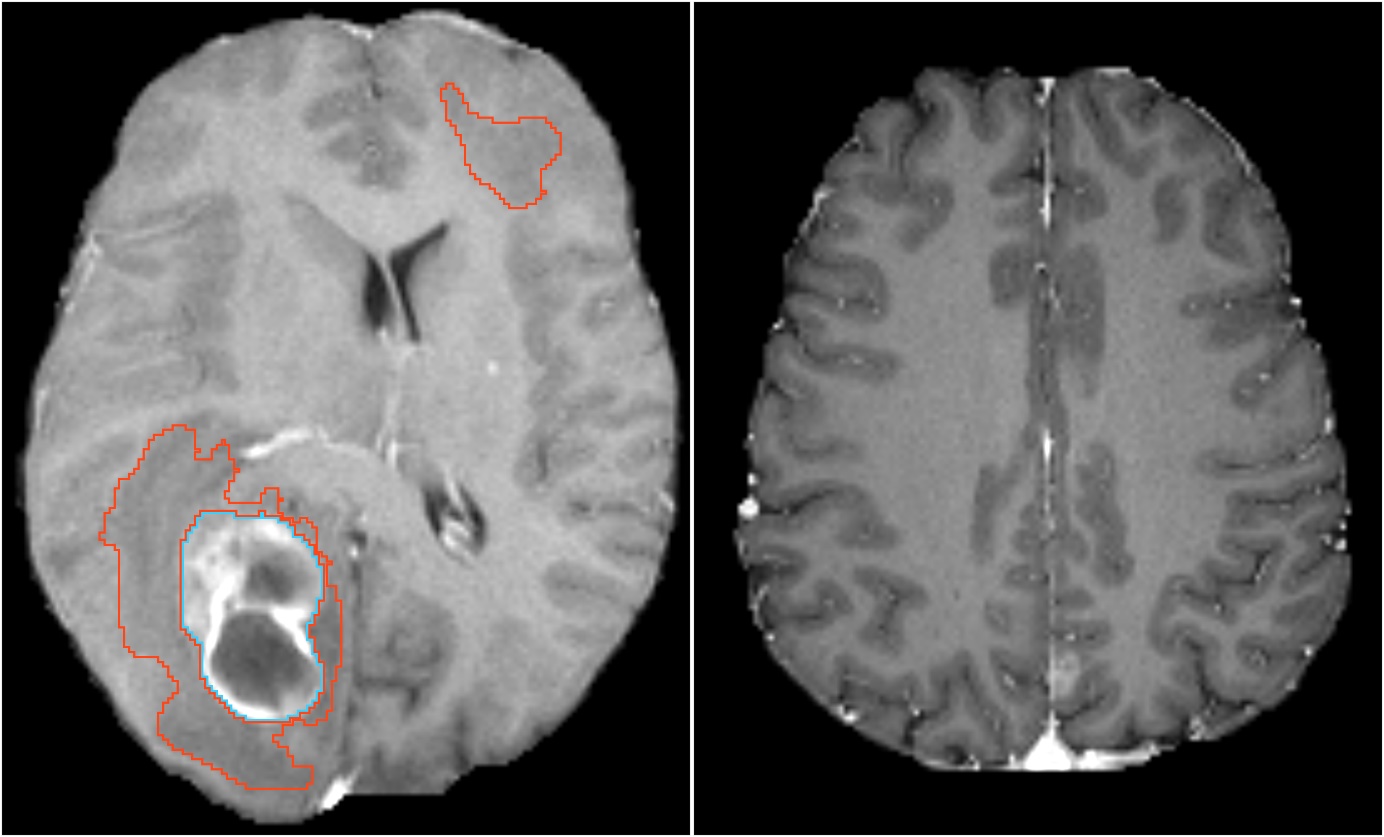

**Supplementary Figure 1: Missed BMs**

In total, five BMs were missed by the T1-CE-only model. One missed BM is shown here in each of the two panels in the T1-CE sequence in axial alignment. On the left, one BM was correctly identified and segmented, whereas a smaller BM (indicated by the blue arrow) was missed. An edema surrounding a third BM (outside this slice) was correctly segmented in the front. On the right, a BM with a connection to the falx was missed.

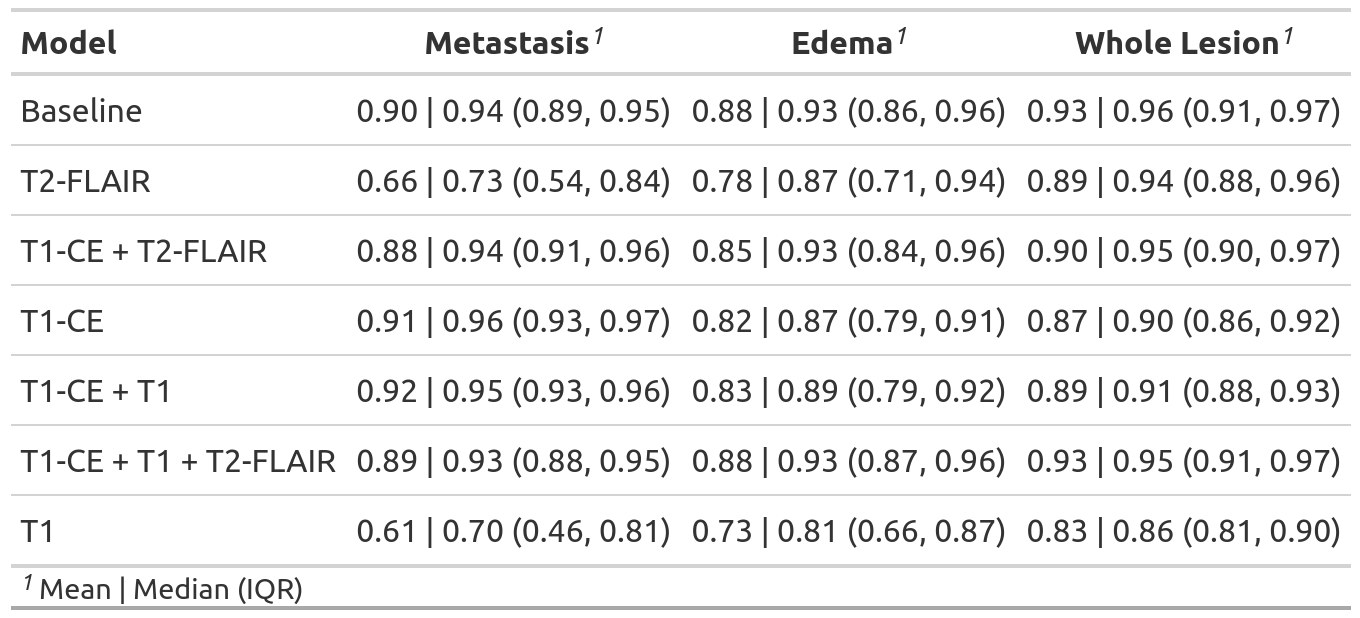

**Supplementary Table 2: Volumetric segmentation performance of all trained models**

The mean and median DSC for the *metastasis*, *edema*, and *whole lesion* (created by combining *metastasis* and *edema* label) label is reported. All 100 patients in our test cohort were analyzed.

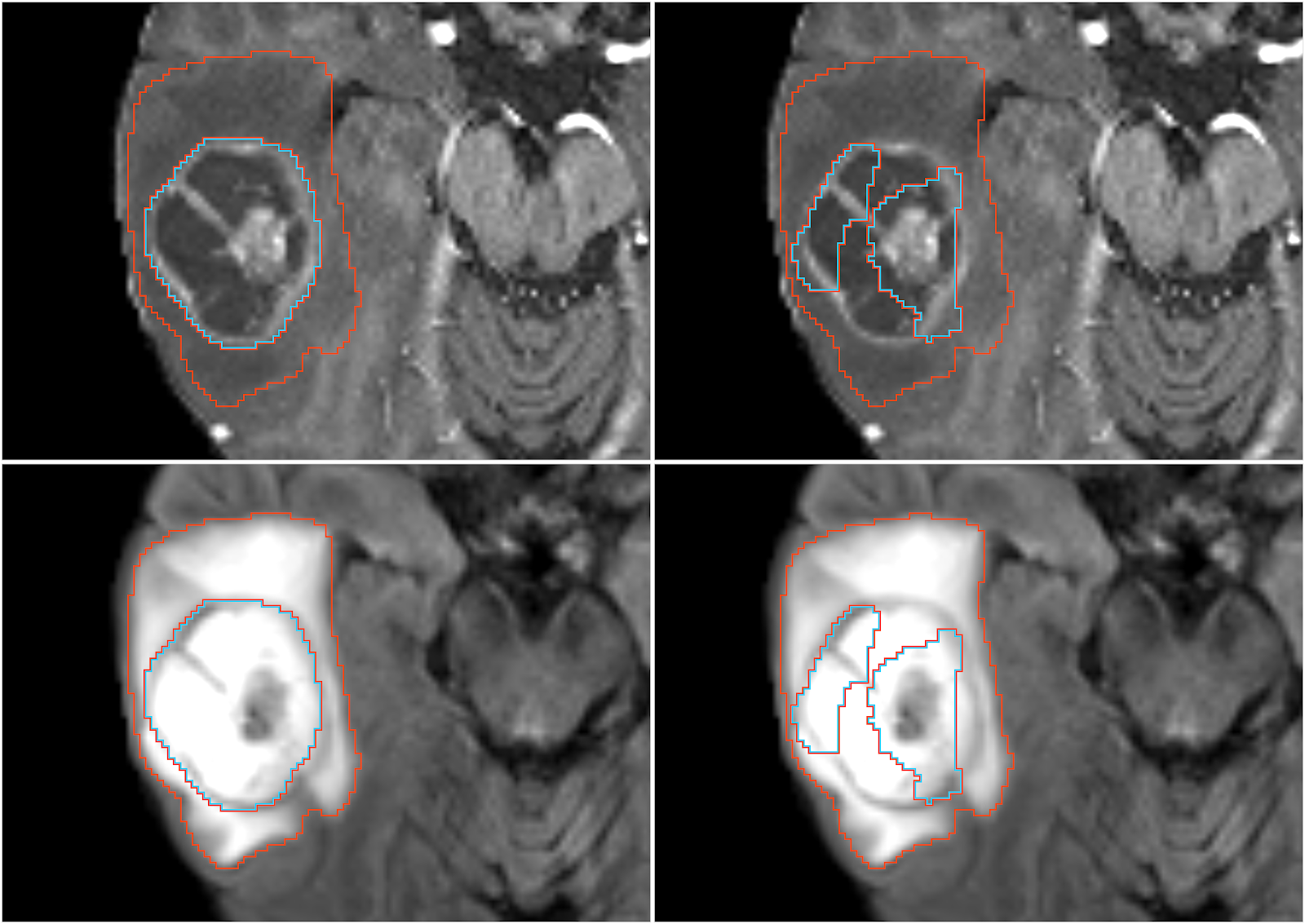
**Supplementary Figure 2: Example patient with misidentified boundary metastasis/edema boundary**

The T1-CE sequence (top) and T2-FLAIR sequence (bottom) are shown in axial orientation. While the T1-CE + T2-FLAIR model on the left correctly labeled the *metastasis* (in blue) and the *edema* (in red), the T2-FLAIR-only model on the right had problems identifying the metastasis/edema boundary. Since the edema usually surrounds the metastasis, the combined *whole lesion* label (*metastasis* + *edema*) was of good quality.

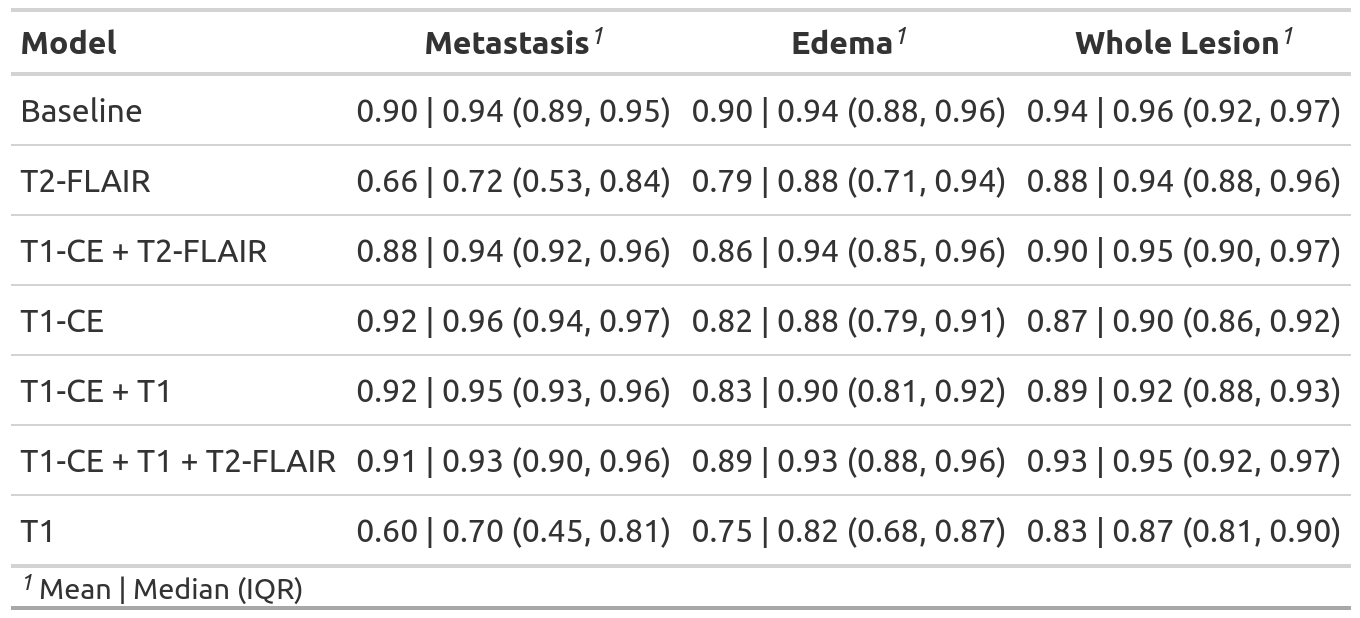

**Supplementary Table 3: Volumetric segmentation performance for patients with all four sequences**

The mean and median DSC for the *metastasis*, *edema*, and *whole lesion* (created by combining *metastasis* and *edema* label) label is reported. In contrast to Supplementary Table 2, patients with synthesized sequences (17 patients with missing T2) were excluded from this analysis. The DSC improved slightly compared to Supplementary Table 2.

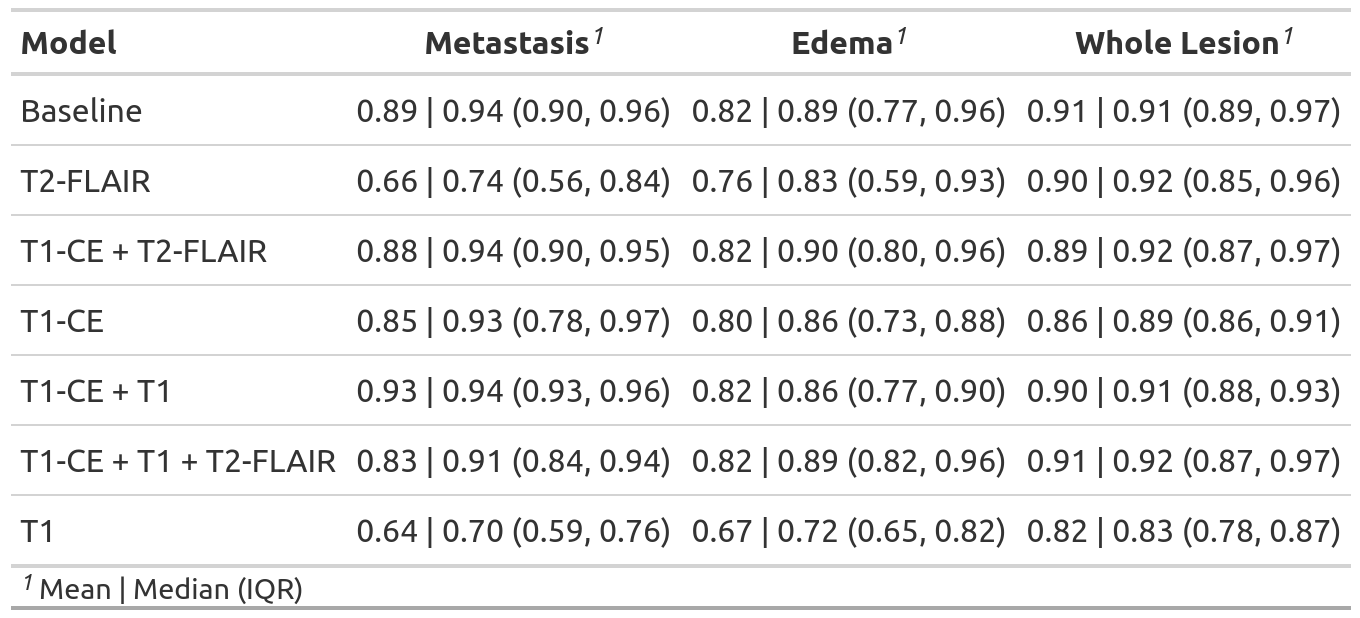
**Supplementary Table 4: Volumetric segmentation performance for the 17 patients with synthetic T2 sequences**

The mean and median DSC for the *metastasis*, *edema*, and *whole lesion* (created by combining *metastasis* and *edema* label) label is reported. The performance of most models was lower compared to Supplementary Table 3, even if T2 was not among the selected sequences. The edema segmentation performance seemed to be especially affected. This may be explained by the overall worse quality of the remaining sequences in patients with a missing T2 sequence.

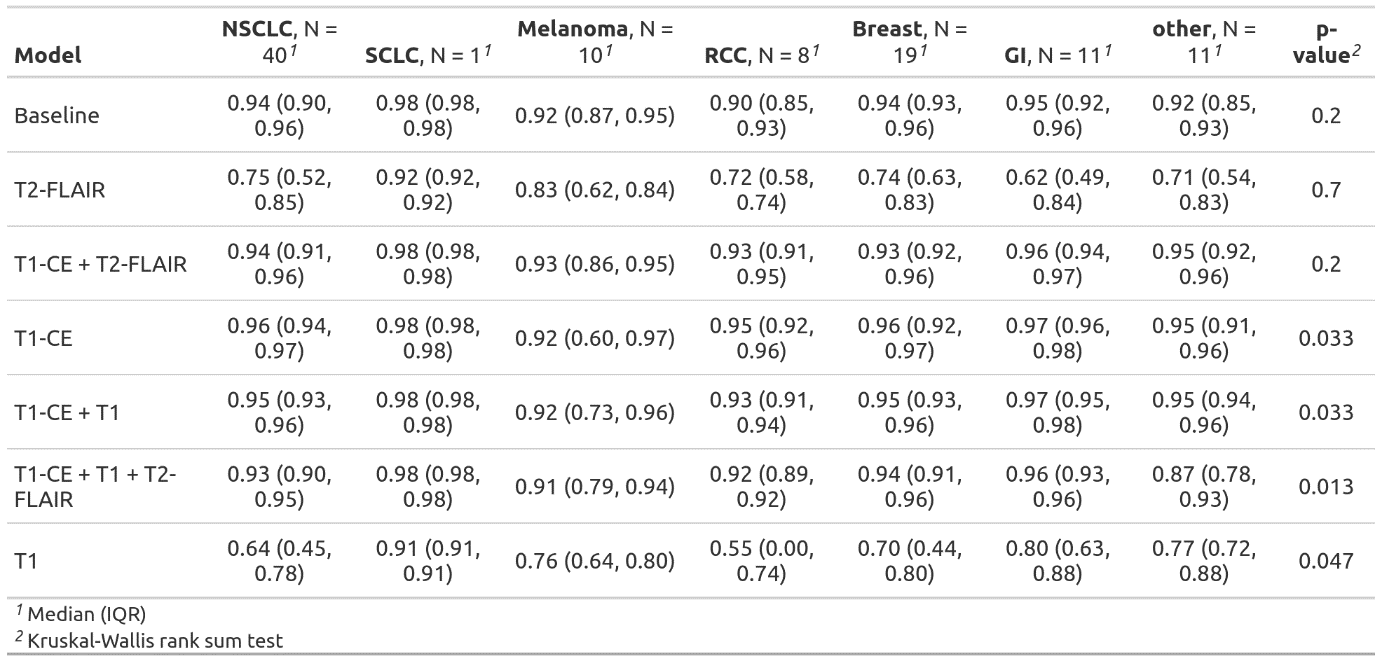
**Supplementary Table 5: DSC for metastasis segmentation depending on histology of primary tumor**

The median DSC for the *metastasis* segmentation is shown depending on the histology of the primary tumor (non-small cell lung carcinoma (NSCLC), small-cell lung carcinoma (SCLC), melanoma, renal cell carcinoma (RCC), breast cancer, gastrointestinal cancer (GI), and others). Small, but significant differences were found in four models.
